# Brain oscillatory modes as a proxy of stroke recovery

**DOI:** 10.1101/2023.02.01.23285324

**Authors:** Sylvain Harquel, Andéol Cadic-Melchior, Takuya Morishita, Lisa Fleury, Martino Ceroni, Pauline Menoud, Julia Brügger, Elena Beanato, Nathalie H. Meyer, Giorgia G. Evangelista, Philip Egger, Dimitri Van de Ville, Olaf Blanke, Silvestro Micera, Bertrand Léger, Jan Adolphsen, Caroline Jagella, Andreas Mühl, Christophe Constantin, Vincent Alvarez, Philippe Vuadens, Jean-Luc Turlan, Christophe Bonvin, Philipp J. Koch, Maximilian J. Wessel, Friedhelm C. Hummel

## Abstract

**Background:** Stroke is the leading cause of long-term disability, making the search for successful rehabilitation treatment one of the most important public health issues. A better understanding of the neural mechanisms underlying impairment and recovery, and the development of associated biomarkers is critical for tailoring treatments with the ultimate goal of maximizing therapeutic outcomes. Here, we studied the longitudinal changes in brain oscillatory modes, linked to GABAergic system activity, and determined their importance for residual upper-limb motor functions and recovery.

**Methods:** Transcranial Magnetic Stimulation (TMS) was combined with multichannel Electroencephalography (EEG) to analyze TMS-induced brain oscillations in a cohort of 66 stroke patients from the acute to the late subacute phase after a stroke.

**Results:** A data-driven parallel factor analysis (PARAFAC) approach to tensor decomposition allowed to detect brain oscillatory modes notably driven by the α frequency band, which evolved longitudinally across stroke stages. Notably, the observed modulations of the α-mode, which is known to be linked with GABAergic system activity, were associated to the extent of motor recovery.

**Conclusions:** Overall, longitudinal evaluation of brain modes provides novel insights into the functional reorganization of brain networks after a stroke and its underlying mechanisms. Notably, we propose that the observed α-mode decrease corresponds to a beneficial disinhibition phase between the early and late subacute stages that fosters structural and functional plasticity and facilitates recovery. Monitoring this phenomenon at the individual patient level will provide critical information for phenotyping patients, developing electrophysiological biomarkers and refining therapies based on personalized excitatory/inhibitory neuromodulation using noninvasive or invasive brain stimulation techniques.

## INTRODUCTION

Stroke is the leading cause of motor disability in the adult population. The exact mechanisms underlying motor impairment and recovery are the targets of a relentless search (*1, 2*), but have yet to be elucidated in detail. Mechanistic knowledge is critical for designing and optimizing future patient-tailored rehabilitation protocols, in contrast to the current “one-size-fits-all” strategy adopted with limited success (*3, 4*). Among the proposed mechanisms, the role of the inhibitory system within the ipsilesional hemisphere has been suggested to be crucially relevant for the course of stroke recovery (*5–7*). Indeed, modulation of inhibition processes can be either beneficial or detrimental for recovery, depending on its direction (increase or decrease) and exact timing. While an increase of the gamma-aminobutyric acid (GABAergic) system activity is beneficial for limiting excitotoxic cell death in the hyper-acute stage, its lasting is detrimental and the presence of a disinhibition phase in later stages is instead favorable for fostering structural plasticity (*7*). However, such phenomena have so far only been examined in animal models or small cohorts of mildly impaired patients (*5, 6, 8–10*). In the present work, we monitored it longitudinally in a cohort of stroke patients, by focusing on brain α oscillations induced by non-invasive cortical stimulation, taking advantage of the coupling between scalp electroencephalography (EEG) and transcranial magnetic stimulation (TMS).

TMS-EEG offers the unique opportunity to directly probe the properties of the neuronal electrophysiological activity (*11*). It inherits both from the causal inference (*12*) and the spatial resolution of TMS (*13*), and from the temporal resolution of EEG, which allows to directly study neuronal oscillations. Since TMS pulses mainly act as a phase reset on neural oscillators (*14, 15*), this tool allows to better characterize the brain dynamics, i.e., evoked neural oscillatory activity, of an area (*16, 17*). Such readouts might be markers of functional reorganization processes that enable motor recovery, especially among the thalamocortical networks to which this technique is sensitive (*17, 18*). Focusing on *evoked* – instead of *induced* - oscillations excludes nonstationary brain activity (*15, 19*), thus eliminating all sources of variability regarding oscillation latency and phase from one stimulation to another (*14, 15*). In a seminal study with healthy subjects, Premoli *et al.* (*20*) explored TMS-induced oscillations over the primary motor cortex (M1) and found specific patterns of oscillations in the α and ß bands. Furthermore, by investigating changes induced by GABAergic drugs, the authors revealed a link between these patterns and the modulation of the GABAergic inhibitory system activity, which is of particular interest in the context of this study.

Recent studies proposed the use of a data-driven approach, the parallel factor analysis (PARAFAC) (*21*), capable of reducing the complexity inherent to TMS-induced oscillations datasets, which are multidimensional (3-5D), into a simpler collection of parsimonious and unique components, or modes (*22, 23*). Applied on TMS-EEG datasets, it allowed to extract three to four brain oscillatory modes in M1 that were physiologically meaningful (*22, 23*).These modes did not overlap in frequency; instead, each was primarily driven by one main oscillatory pattern in the θ, α or β band, each related to a specific mechanism. The study of the mode associated with late TMS-induced α oscillations enables the monitoring of the evolution of inhibitory processes throughout the course of post-stroke motor recovery, since variations within this band are linked with functional inhibitory processes (*24–29*) mediated by the GABAergic system (*20*).

In the present study, we sought to elucidate the mechanisms underlying motor impairment and recovery after stroke by applying this powerful method in a large stroke cohort evaluated longitudinally from the acute to the late subacute stage, in the framework of the TiMeS project (*30*). The present work complements the study of TMS-evoked responses (*31*), the results of which will be put into perspective with those of the present study. Here, we specifically studied the changes in brain oscillatory modes unveiled from TMS-induced oscillations and determined their importance for residual motor functions and recovery.

## MATERIALS AND METHODS

This work is part of the TiMeS project, a detailed description of which is provided in our previous work (*30, 31*), and in the *Supplemental Material* (*SM*).

### Study design

TMS-induced oscillations and behavior were assessed at three different time points, one week (referred to here as “acute stage”, A), three weeks (“early subacute stage”, ESA) and three months poststroke (“late subacute stage”, LSA, Fig. 1A), on the same cohort of 66 patients previously described in (*31*) (see *SM*). Additionally, in order to validate and replicate the PARAFAC approach on TMS-EEG data, 19 healthy young adults (age: 26.9 ± 2.9 years, 9 females) and 15 healthy older adults (aged-matched with patients, age: 67 ± 5 years, 11 females) were recruited and underwent a single TMS-EEG recording session. The study was conducted in accordance with the Declaration of Helsinki and was approved by Cantonal Ethics Committee Vaud, Switzerland (project number: 2018-01355). Written informed consent was obtained from all participants.

**Fig. 1:**
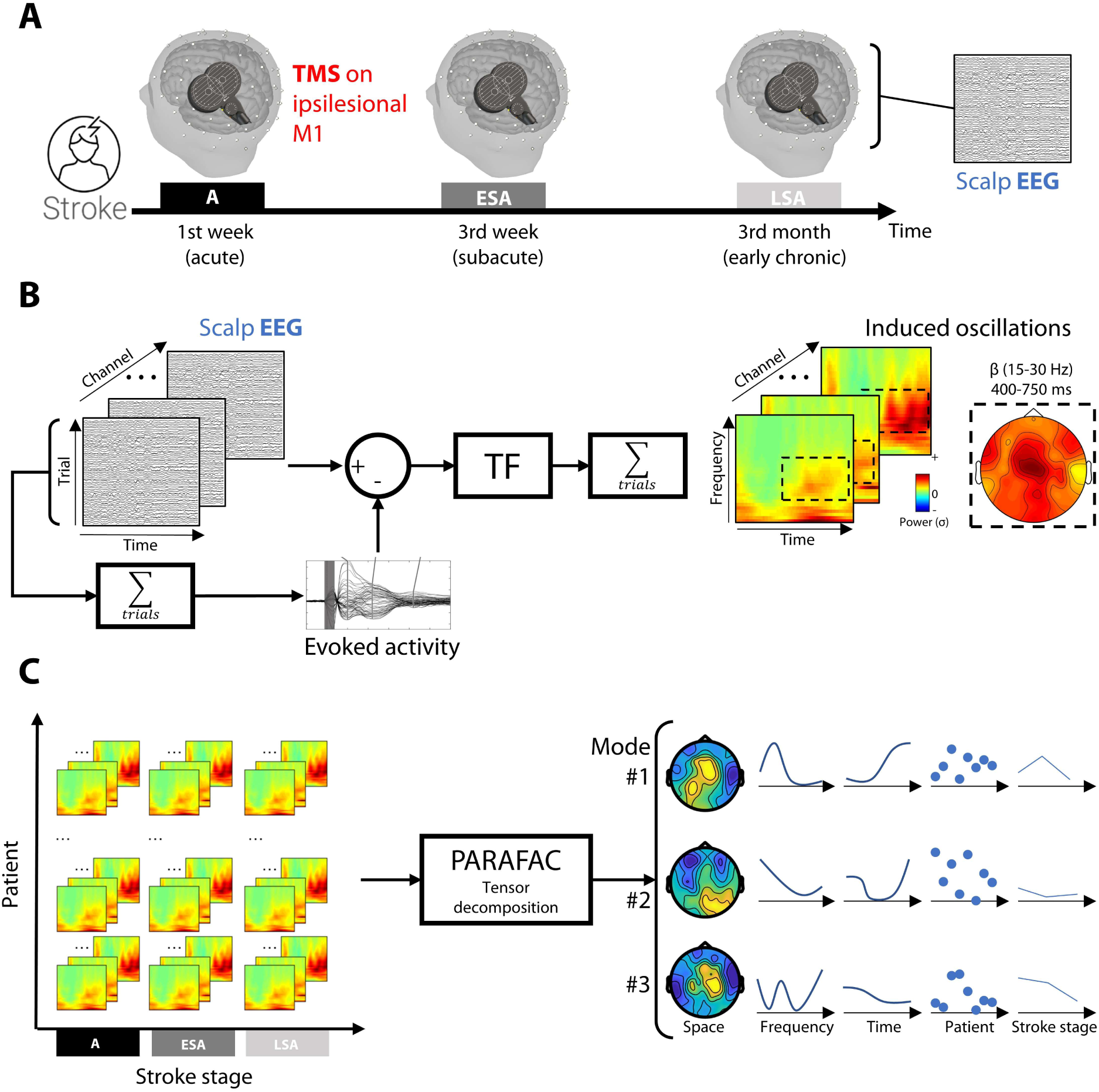
Protocol design and main data processing pipeline. (**A**). Protocol design including 3 TMS-EEG sessions (referred to as the acute, early subacute and late subacute stages, respectively). (**B**). Signal processing pipeline for computing induced oscillation maps. For each patient and channel, the evoked activity was removed from the clean signal prior to the time-frequency (TF) transform. Each TF map was z transformed before averaging across trials. (**C**). Tensor decomposition using the PARAFAC approach. Induced oscillation maps were compiled into a tensor, with the patient and stroke stages as the 4^th^ and 5^th^ dimensions, respectively, and decomposed using the PARAFAC algorithm. This decomposition led to several components, or modes, whose weights are represented in the space (topography), frequency, time, patient and stroke-stage dimensions, from left to right.

### TMS-EEG analysis

The analysis methodology used in this study was adapted from Tangwiriyasakul *et al.* (*22*), in which the PARAFAC tensor decomposition approach was applied to the preprocessed TMS-EEG data of our previous work (*31*).

#### Induced oscillations

Induced oscillations were computed in MATLAB using the Fieldtrip toolbox (*32*) (Fig. 1B). First, the TMS-evoked potentials (TEPs) were computed by averaging all trials and were then individually subtracted from the signal in each trial to filter out evoked activity (*33*). Then, the time-frequency (TF) map of each corrected trial was computed using a multitapers approach. The signal from the -500 to +1,000 ms time window (10-ms step) was convoluted with Hanning tapers ranging from 7 to 40 Hz (1-Hz step), with a width of 3.5 cycles per window. For each electrode and trial, the resulting power time series was normalized using the z-score against baseline (−200 to -50 ms), before being averaged across trials to obtain the final TF maps. Prior to tensor definition, all the TF maps were flipped as needed so that the ipsilesional hemisphere was defined as the left for all patients.

#### PARAFAC tensor decomposition

Five different tensors were constructed as follows: the first tensor focused on the A stage (4D: electrode × frequency × time × patient), while the second to the fifth tensors gathered the induced oscillations across stroke stages (A vs. ESA stage, A vs. LSA stage, ESA vs. LSA stage, and from A to LSA stage; 5D: electrode × frequency × time × patient × stroke stage) (Fig. 1C). For each tensor, the TF maps were cropped between +40 and +750 ms to prevent missing values from boundary effects, resulting in a size of 62 × 34 × 70 for the first three dimensions. The fourth-dimension size was equal to the number of patients included at the A stage (60), the A and ESA or LSA stages (43 or 33 respectively), the ESA and LSA stages (30), and all stages (27). Tensor decomposition was performed using the N-way toolbox (Rasmus Bro, 2024; https://www.mathworks.com/matlabcentral/fileexchange/1088-the-n-way-toolbox) using the non-negativity constraint to all dimensions.

### Statistical analysis

Differences between stroke stages when decomposing the 5D tensors were assessed using the same permutation-based approach proposed in (*22*) (see *SM*). The link between patient weights (data in the 4^th^ dimension) within the extracted modes and motor scores for each stroke stage and change ratios between stroke stages (see *SM*) was explored using the Bayesian equivalent of nonparametric Kendall correlation and partial correlation testing using JASP software (JASP Team – 2023). The weights of each extracted mode were compared with the initial motor scores in the A stage and to the change ratio between stroke stages. The default values proposed within the JASP framework were used to keep the priors regarding effect sizes relatively large. Correlation values were reported using the 95% confidence interval of Kendall’s τ, while the statistical evidence of the tests was reported using Bayes factors (BF_10_) and the cutoff values defined by Jeffreys, 1998 (*34*) for interpretation.

## RESULTS

### Brain oscillatory modes in acute stroke patients

Fig. 2 shows the induced oscillatory modes obtained after the 4D tensor decomposition in A patients. Overall, the decomposition allowed us to identify three modes that were similar to those found on healthy young and older participants (see *SM* & Fig. S1), and to those previously reported in the literature (*22, 23*). The first mode was mainly driven by low frequencies, and was characterized by the typical 1/f spectral trend of aperiodic activity, mostly located over the stimulation site. The second mode converged on the late parieto-occipital α waves (8-10 Hz peak for A patients and healthy controls) that emerged over time starting 150-200 ms after stimulation. The third mode mainly focused on central sensorimotor β waves. The exact peak frequencies tended to differ among the three groups: A patients presented one peak at 15 Hz, while healthy young adults showed two peaks at 7 and 21 Hz, and older adults showed one peak at 19 Hz (see Fig. S1).

**Fig. 2:**
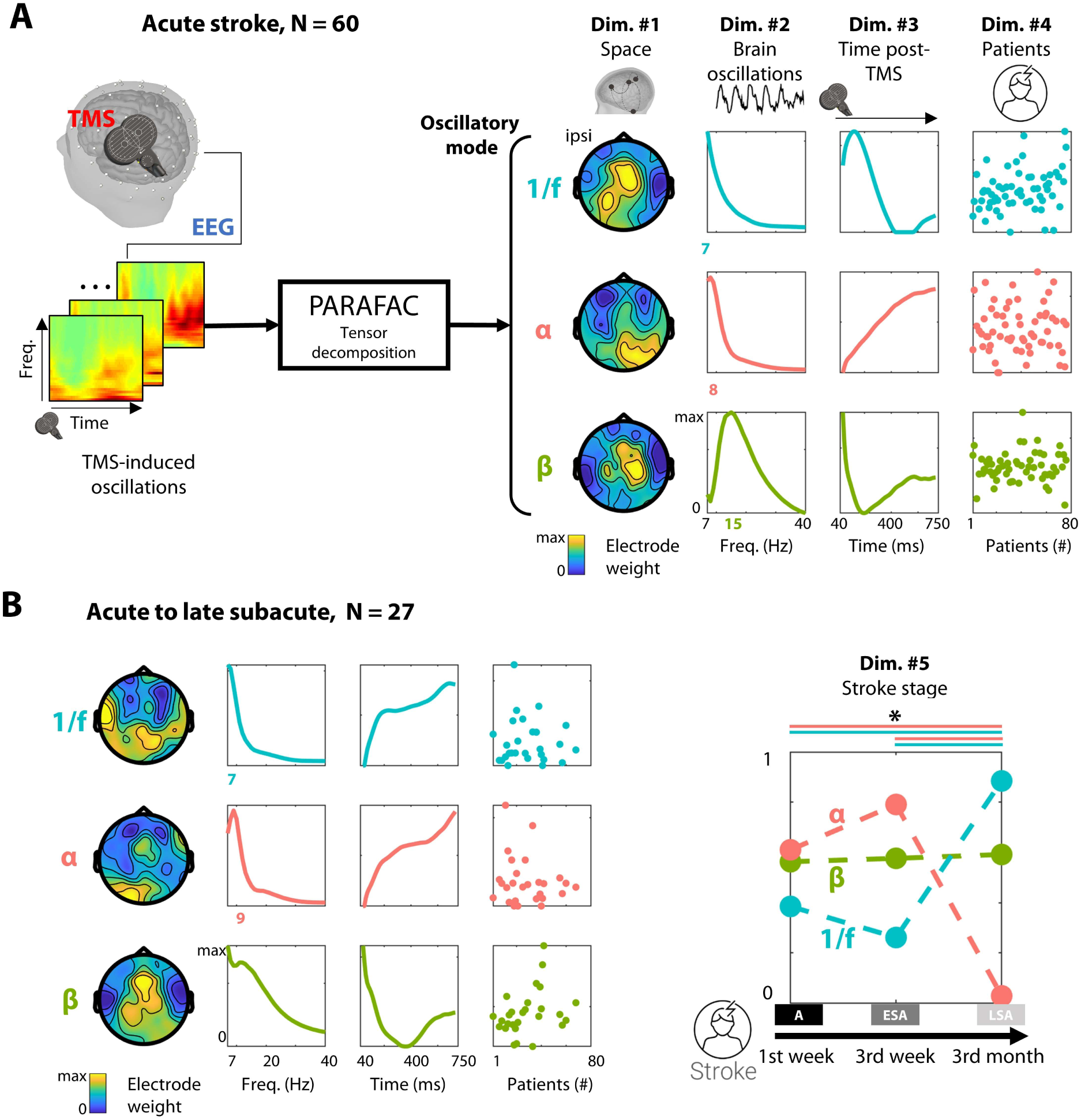
Brain oscillatory modes in acute stroke patients and their modulation towards subacute stages. **(A)** PARAFAC decomposition of the 4D tensor of the TMS-induced oscillations in acute stroke patients. Modes are sorted by row according to their main frequency peak, from low frequencies (1/f spectral trend, blue) and α (8 Hz, red) to β (15 Hz, green) frequency bands (top to bottom). Each column depicts the relative weights (from 0 to maximum) of each mode in the space, frequency, time and patient dimensions (from left to right). The mode frequency peak is highlighted in color on the y-axis. Data were flipped for patients whose lesion was located on the right hemisphere so that the ipsilesional side was the left side. (**B**) PARAFAC decomposition of the longitudinal 5D tensor in all patients. The modes are represented identically, except for the addition of a fifth dimension representing the stroke stage (right). The asterisk and colored lines indicate significant effects of the pairwise comparisons of the stroke stages within the 1/f (blue lines) and α (red lines) modes for the acute (A) vs. late subacute (LSA) stages and the early subacute (ESA) vs. late subacute (LSA) stages (permutation test, p_Bonf_ < 0.05, see *Statistical analysis*).

### Evolution of brain oscillatory modes over the time course of recovery

The results of the decomposition of the 5D tensors including all stroke stages (A, ESA and LSA) are presented in Fig. 2B. Interestingly, the mode weights were significantly modulated across stroke stages (p_Bonf_ < 0.05, see Fig. S2 for detailed results). This change was specific to 1/f and α-band modes that significantly differed from the A and the ESA stages to the LSA stage with opposite directions. While the relative weight of the α-band mode was stable at the ESA stage before significantly decreasing at the LSA stage, the aperiodic mode modulated in the opposite direction with a significant increase at the LSA stage. Overall, the changes were much stronger at the LSA stage, which was confirmed in larger groups of patients by pairwise comparisons between stroke stages: no change was found for any of the modes when comparing the A to the ESA stage (Fig. 3A), whereas a strong increase and decrease in the 1/f and α-band modes, respectively, was found toward the LSA stage (p_Bonf_ < 0.05, Fig. 3B&C). Finally, no significant modulation of the β-band mode through stroke stages was found in any of the tensor decompositions.

**Fig. 3:**
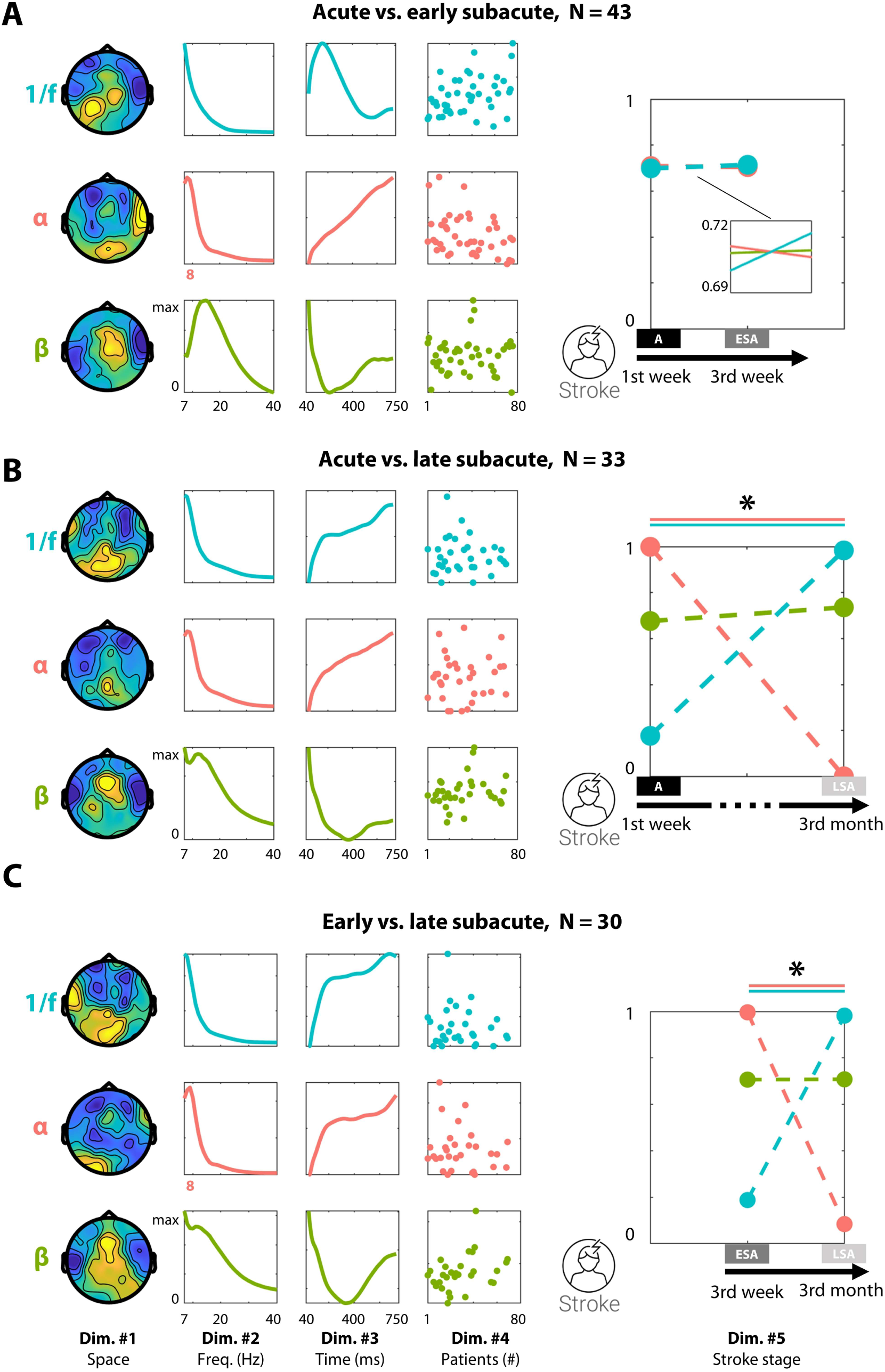
Evolution of brain oscillatory modes from acute to subacute stages. (**A**) PARAFAC decomposition of the 5D tensor in all patients. The modes are sorted by peak frequency, from 1/f (blue) and α (red) to β (green) bands. Each column depicts the relative weights (from 0 to maximum) of each mode in the space, frequency, time, patient and stroke-stage dimensions (from left to right), limited here to A and ESA. (**B and C**). The results of the same decomposition, run separately on the A and LSA stages **(B)** and on the ESA and LSA stages **(C)**. The asterisk and colored lines indicate significant effects of the pairwise comparisons of the stroke stages (permutation test, p_Bonf_ < 0.05, see *Statistical analysis*).

### Modulation of brain oscillatory modes as a proxy of motor recovery

We further explored the modulation across stroke stages by distinguishing patients who actually recovered along the evaluated stroke stages (recovering group) from patients who maintained stable motor functions/impairment since their inclusion at the A stage (stable group, for group definitions please see *SM*). The two groups drastically diverged regarding the modulation across stroke stages. These data indicate that the previously observed modulations were mainly driven by the recovering group, in which the same significant effects were observed between ESA and LSA stages (p_Bonf_ < 0.05; Fig. 4A, right), whereas no change was noticeable within the stable group between stroke stages (Fig. 4A, left).

**Fig. 4.**
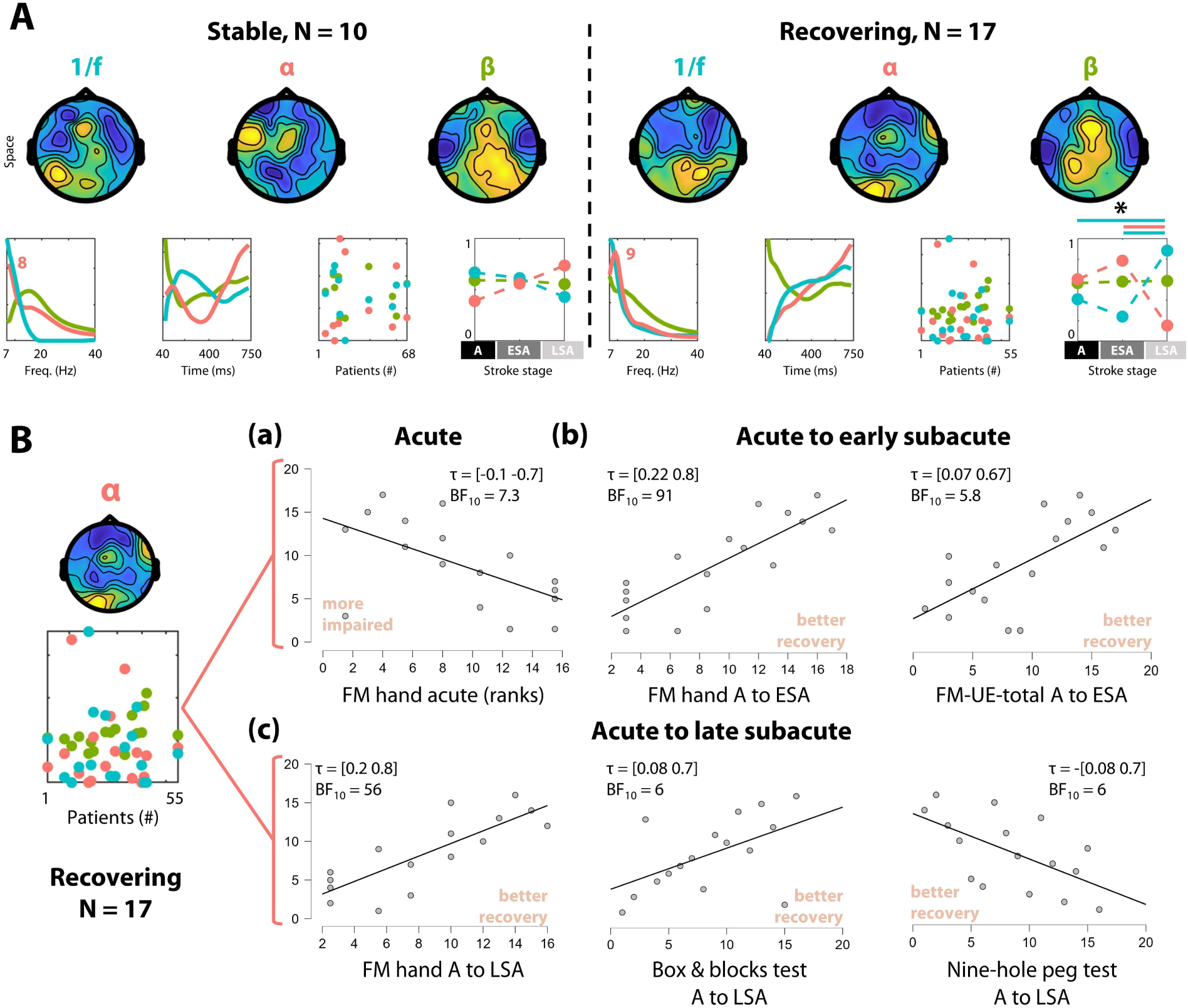
Links among brain oscillatory modes, motor impairment and motor recovery. (**A**). PARAFAC decomposition of the 5D tensor in stable (left) and recovering (right) patients. (**B**) Association between the α-band mode and (a) motor impairment in the acute stage, and (b-c) motor recovery towards the early (b) and late (c) subacute stages. The Kendall rank correlation coefficient (τ) and Bayesian factors (BF_10_) are indicated for each comparison, and all data are plotted according to their rank.

All tensor decompositions showed variability in the weights of modes among patients, i.e., within the 4^th^ dimension. We next aimed to explain the variability among patients in terms of impairment, function and recovery, by linking this variability with the different modes and their changes (Fig. 4B). No statistical evidence was found for either a link or an absence of links between 1/f and β-band modes on motor scores (all 1/3 < BF_10_ < 3) in any of the tested patient cohorts. However, moderate to strong statistical evidence was found for a link between the α-band mode and motor scores in the A stage and its evolution across the ESA and LSA stages in the recovering group. First, patients who exhibited a stronger weight associated with this α-band mode were more impaired in the A stage (Fig. 4B(a)), with lower Fugl-Meyer (FM) hand scores (τ = [-0.09 -0.7], BF_10_ = 7.3). Broader associations with the α-mode were found in better recovering patients between the A and ESA stages (Fig. 4B(b)), as revealed by a stronger change ratio between the A and ESA stages in the FM hand (τ = [0.22 0.8], BF_10_ = 91; BF_10_ = 6.5 when controlling for the significant association with the initial level of impairment), wrist (τ = [0.05 0.65], BF_10_ = 3.9), and UE-total scores (τ = [0.07 0.67], BF_10_ = 5.8) as well as the maximum fist force (τ = [0.07 0.67], BF_10_ = 5.9). The stronger the patients were associated to this α-band mode, the better they improved at the ESA stage. Finally, a similar association was found in the longer term, with stronger change ratios between the A and LSA stages in scores on the FM hand (τ = [0.2 0.8], BF_10_ = 56; BF_10_ = 12 when controlling for the initial level of impairment), BnB (τ = [0.08 0.7], BF_10_ = 6.0) and nine-hole peg tests (τ = [-0.08 -0.7], BF_10_ = 6.0) (Fig. 4B(c)), indicating that greater changes in the α-band mode were associated with larger improvements in motor functions and thus larger reductions in impairment.

## DISCUSSION

In the present study, we report longitudinal changes in induced oscillatory activity associated with functional motor recovery in a cohort of stroke patients. The results, together with other current work (*31*), highlight dynamic changes in correlates of inhibitory (GABAergic) activity towards a recovery-supporting disinhibition towards the late subacute stage (for a schematic summary please see Fig. 5).

**Fig. 5.**
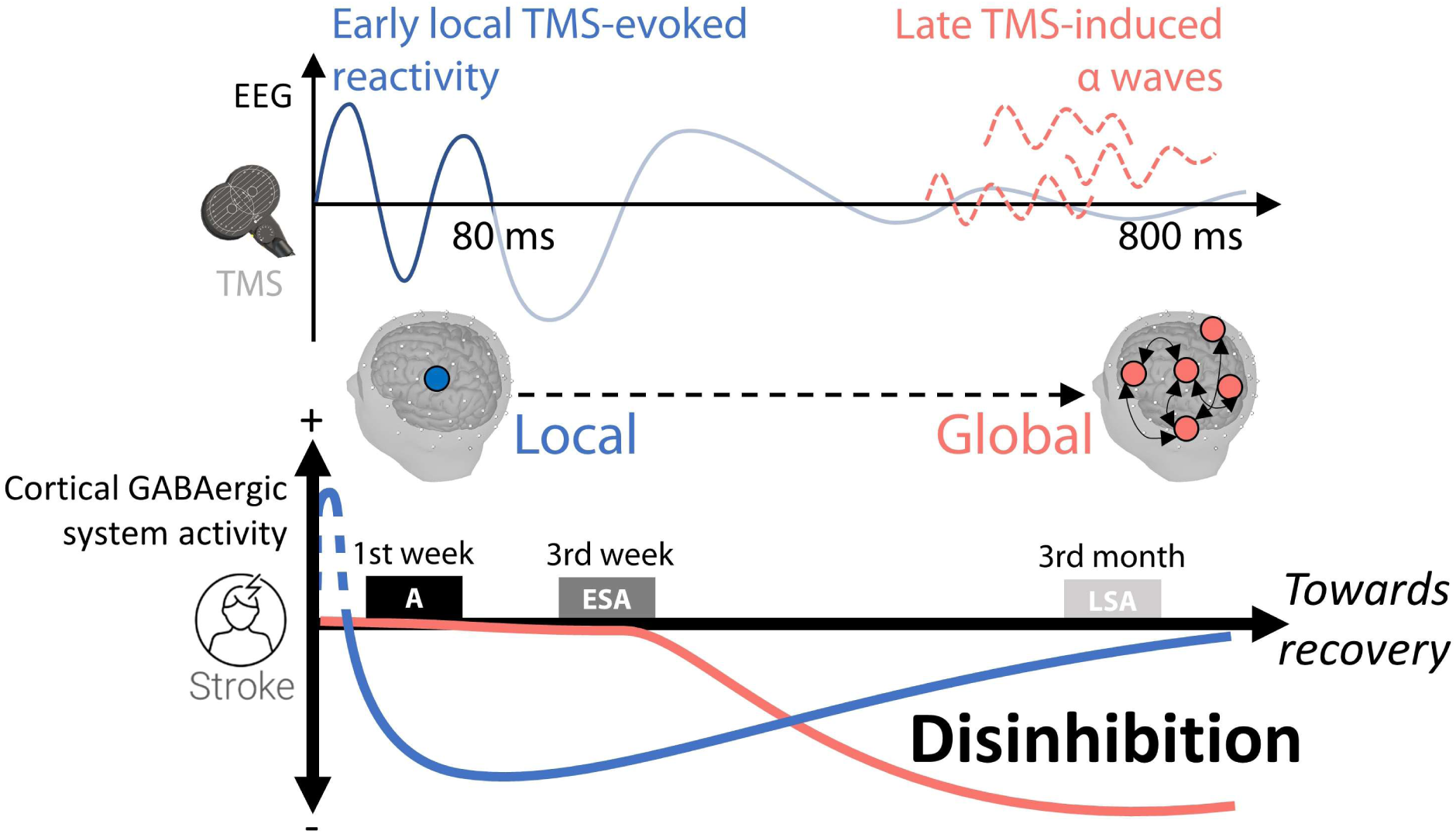
Time course of GABAergic inhibition and its relationship with recovery. The changes in early TMS-evoked reactivity (as demonstrated in Harquel *et al.*, 2024(*31*)) and in late TMS-induced α waves, both most likely correlates of a decrease in GABAergic activity at different spatial scales, represent recovery-facilitating functional disinhibition after the detrimental hyper-inhibitory period in the hyper-acute stage after the stroke. Based on the present findings, this disinhibition occurs in two phases: first locally within the ipsilesional motor cortex in the acute to subacute stage, and then more broadly towards the late subacute stage. Such disinhibition fosters structural and functional plasticity that support motor recovery (*41, 42, 48*).

The evolution of TMS-induced oscillations might be a relevant proxy of functional reorganization and its underlying mechanisms. The longitudinal increase and decrease observed in the 1/f spectral trend and α-band modes respectively, together with the absence of any significant change within the β-band mode, underline the importance of the evolution of the excitatory/inhibitory balance across stroke stages (Fig. 2B & 3). On the one hand, the 1/f spectral trend is an ubiquitous feature of scalp EEG (*35*) reflecting non-oscillatory aperiodic neural activity (*36*). Whereas less studied than neural oscillations, the observed inter- and intra-subject variability of aperiodic activity has been shown to relate notably to aging (*37*) and conscious states (*38*). Recent work suggested that the spectral trend is modulated by several physiological mechanisms, such as synaptic timescale and excitatory/inhibitory balance (*39, 40*). Accordingly, the observed positive modulation trough time post-stroke may serve as a potential correlate of changes within the excitatory/inhibitory balance, which are recognized as crucial for recovery (*7*). However, there are current limitations in our understanding of the precise origin and nature of the aperiodic activity (*39*). In addition, we are examining a TMS-induced phenomenon (rather than a resting-state or a task-related one, as in the previously cited literature). Hence, we cannot determine the precise direction of these changes (regulation or alteration) on the basis of aperiodic component modulation alone. On the other hand, TMS-induced α- and β-band activity has been linked with the GABAergic (*20*) and glutamatergic (*23*) systems respectively. Therefore, our results rather pointed to a dominantly GABAergic activity changing compared to glutamatergic activity, as we observed variations in α-but not in β-band. This finding points to the existence of a beneficial disinhibition phase, especially in the recovering patient group (Fig. 4).

The α-band modulations found here can thus be considered a proxy of the dynamic evolution of the intracortical inhibitory system (*20*), which has been suggested to sustain motor recovery ((*8, 41*). Immediately after stroke, during the hyperacute phase, hyperinhibition of the perilesional cortical areas prevents additional tissue damage due to ischemia-induced excitotoxicity (*42–44*). However, the persistence of hyperinhibition over time was correlated with worse motor outcomes (*6, 8*), and pharmacological reduction in GABAergic inhibition led to better recovery in animal models (*6, 45, 46*). Further evidence has confirmed this last point by linking better motor recovery with a period of plasticity driven by molecular changes, such as cellular excitability (*47*), or by sustained disinhibition during the first weeks poststroke. Moreover, the existence of this beneficial disinhibition stage is also confirmed in the present cohort of patients when specifically studying cortical reactivity (*31*). Our results showed that an abnormally stronger reactivity, supported by intracortical motor disinhibition assessed via paired-pulse TMS, was linked with better motor outcome. Overall, this disinhibition has been suggested to promote functional reorganization within the lesioned hemisphere (*41, 42, 48*). Consistent with this suggestion, the decrease in the α-band mode in the recovering patient group is most likely a correlate of the disinhibition towards the late subacute stage (three weeks to three months poststroke), which supports the recovery process (please see Fig. 5 for a schematic).

The time frame of this phenomenon is somewhat delayed compared with our findings regarding brain reactivity on the present cohort (*31*) and the previous findings of Liuzzi *et al.* (*8*), in which disinhibition occurred from the first days to up to three weeks after stroke onset. The slight differences in the precise timing of this effect might stem from the brain areas and networks represented by the specific measures and temporal windows from which they are extracted. The electrical activation of neuronal populations subsequent to the TMS pulse is first spatially restricted to the targeted cortical site, before propagating along white matter pathways to reach distant cortical and subcortical sites (see, *e.g.,* (*49*)). Then, while the short-interval intracortical inhibition protocol used in our study (*31*) and by Liuzzi *et al.* (*8*) allows measurement of the intracortical GABAergic activity locally within the motor cortex, the late induced α oscillations in the present work are linked with inhibitory activity at a more global scale, i.e., engaged in higher-order processes within larger-scale brain networks (Fig. 5). The bottom-up propagation of TMS has been well established (*50*). While the stimulation is first triggering local activity within local nodes, it is then propagating to connected brain networks including hubs, such propagation being reflected in the temporality of the EEG signal, from early to late activity(*49, 50*). Thus, the disinhibition phenomenon might first occur locally within the lesioned motor cortex in the acute to subacute stage before spreading to larger brain networks including hubs to promote functional plasticity more broadly and thus support more complex motor functions in later stages, as revealed here by the later association with improvement in complex motor tasks such as the BnB or 9HP.

### Limitations

Recent guidelines about TMS-EEG acquisition and data processing were followed in this work, with the exception of the presence of a realistic sham stimulation condition in order to assess the influence of peripheral evoked potentials, due to the multisensory features of TMS, on the recorded EEG signal (*51, 52*). However, since the present work is based on TMS-induced oscillations, which are computed after removing such evoked components from the signal, and on a longitudinal analysis approach, the lack of a sham stimulation condition does not relevantly affect the interpretation of the results. In addition, the distribution of the patient cohort with respect to the severity of motor impairment in the acute stage was rather on the moderately to mildly impaired side. Additional analyses of more heterogeneous groups in the future will help to refine the present conclusions regarding the link between changes in brain oscillatory modes and motor recovery.

### Conclusions

In summary, examining a larger cohort of stroke patients over time using TMS-induced activity provided a better understanding of the neural mechanisms linked to motor recovery. The present results support cortical disinhibition with different topographies and temporal course as an important underlying mechanism driving motor recovery between the early and late subacute stages. The acquired knowledge might pave the way for developing novel biomarkers to determine and predict stroke recovery and to personalize innovative therapies based on modulation of brain oscillatory activity through noninvasive or invasive brain stimulation technologies.

## ACKNOWLEGMENTS

We thank the MRI and neuromodulation facilities of the Human Neuroscience Platform of the Fondation Campus Biotech Geneva and the Neuroimaging Center of the Sion Hospital and the Center for Biomedical Imaging, a Swiss research center of excellence, for their expertise and access to their facilities.

## Data Availability

The data related to this article are available upon reasonable request from the corresponding author.

