## Supplemental material for "Brain oscillatory modes as a proxy of stroke recovery"

### MATERIALS AND METHODS

#### Inclusion and exclusion criteria

The inclusion criteria were as follows: older than 18 years of age, no contraindications for MRI or TMS, and (for the patient group) motor deficits of the upper limb. The exclusion criteria included cognitive inability to provide informed consent, history of seizures, pregnancy, severe neuropsychiatric or medical diseases, regular use of narcotic drugs or medication that significantly interact with TMS, implanted medical electronic devices or ferromagnetic metal implants incompatible with MRI or TMS, and the request not to be informed in case of incidental findings.

#### Behavioral data

At each time point, the bilateral motor capability and impairment of patients were assessed using (i) the Fugl-Meyer Assessment (FM) of the upper extremity (FM-UE-total, max 60 points without reflexes), of the upper extremity (FM-UE, max 30 points), of the hand and of the wrist, (ii) the maximum fist, key and pinch force, (iii) the box and blocks test (BnB), and (iv) the nine-hole peg test. The maximum grip forces were assessed in three trials using a JAMAR® hydraulic hand dynamometer. The ratio between the affected and the nonaffected hand was used as the primary outcome for the grip forces and scores on the box and blocks and nine-hole peg tests. Change ratios between time points were computed for each motor measurement $x$ as follows (e.g., A vs. ESA): $(x_{ESA} - x_A)/x_A$. Patients were classified as "recovering" if they exhibited any improvement in FM-UE scores (1 point minimum).

### TMS-EEG acquisition

The details of the acquisition parameters can be found in our previous paper *(31)* that analyzed TMS-evoked potentials. In brief, 64-channel TMS-compatible EEG (BrainAmp DC amplifiers, Brain Products GmbH, Germany) was recorded concurrently with the neuronavigated (Localite GmbH, Germany) stimulation of the ipsilesional motor cortex over the first dorsal interosseous (FDI) motor hotspot using an MC-B70 coil connected to a MagPro X100 stimulator (MagVenture A/S, Denmark). A total of 180 suprathreshold single-pulse stimulations were delivered. If no motor activity was evoked from the lesioned hemisphere, the stimulation parameters were tuned to the contralesional hemisphere. DC and low-pass (1 kHz) filtered EEG was sampled at 5 kHz, and the electrode impedance level was kept below 5 kΩ. During the stimulation, the patient was instructed to remain still while staring at a fixation cross and listening to white noise through noise-canceling earphones to limit eye movements and the influence of the TMS click sound on the EEG signal.

### TMS-EEG data preprocessing

TMS-EEG data were analyzed with MATLAB (The MathWorks, USA) and preprocessed using the EEGLAB and TESA toolboxes. The clean datasets analyzed in this study were identical to those in our previously published paper *(31)*. In short, raw TMS-EEG data were preprocessed using a double independent component analysis (ICA) to remove components linked to pulse, muscle, decay and ocular artifacts from the data. The preprocessed dataset resulted in an average of 149 ± 24 cleaned trials epoched from -500 to +1,000 ms around the TMS pulse and filtered between 1 and 80 Hz.

### Tensor sizes

A total of 60, 43, 33, 30 and 27 patients were included at the A stage, the A and ESA stages, the A and LSA stages, the ESA and LSA stages, and all stages, respectively; thus, the final sizes of the five tensors were as follows: first tensor (A stage only), 62 × 34 × 70 × 60 (8,853,600 datapoints); second tensor (A vs. ESA stage), 62 × 34 × 70 × 43 × 2 (12,690,160 datapoints); third tensor (A vs. LSA stage), 62 × 34 × 70 × 33 × 2 (9,738,960 datapoints); fourth tensor (ESA vs. LSA stage), 62 × 34 × 70 × 30 × 2 (8,853,600 datapoints); and fifth tensor (A to LSA; all stages), 62 × 34 × 70 × 27 × 3 (11,952,360 datapoints). The link between brain oscillatory modes and motor recovery was further inspected by splitting the last 5D tensor into two parts: the first subtensor comprised only stable patients, while the second comprised recovering patients. Regarding healthy adults, the final tensor of the healthy young adults group was 62 × 34 × 70 × 19 (2,803,640 datapoints) and that for the healthy older adults group was 62 × 34 × 70 × 15 (2,213,400 datapoints).

**Statistical analysis**

Differences between stroke stages when decomposing the 5D tensors were assessed using the same permutation-based approach proposed in *(22)*. 1,000 surrogate tensors were obtained by permuting data from the $4^{th}$ (patient) and $5^{th}$ (stroke stage) dimensions while keeping the data structure unchanged over the 3 first dimensions. A PARAFAC decomposition was then performed on each surrogate tensor using the old loadings of the original data decomposition in the first three dimensions. The corresponding mean differences between stroke stages were gathered across all decompositions to form the surrogate data distributions. Differences between stroke stages within each brain mode (1/f, α and β modes) in the original tensors were considered significant if greater or lower than 2.5% of these surrogate distributions ($p < 0.05$, two-sided). This α level was Bonferroni corrected in order to account for multiple testing (1 or 3 differences × 3 modes = 3 or

9, for tensors including 2 or 3 stroke stages respectively), which led to a corrected threshold of 0.83 % or 0.25 % respectively (referred as $p_{Bonf} < 0.05$ in the text).

### RESULTS

#### Study sample

A total of seventy-six stroke patients were initially enrolled in the study following their admission to the Cantonal Hospital in Sion, Switzerland. Of these, 66 patients (mean age: 68.2±13.2 years; 18 females) were included in the analysis, having undergone TMS-EEG recordings in at least one session (refer to Figure 2A for the screening flow chart, and Table 1 and Figure 2B for patient characteristics, in our previously published paper *(31)*). To identify factors associated with recovery, a subgroup of patients exhibiting motor improvement was designated as the recovering group (N=40). This recovery was quantified by an increase of at least one point in the Fugl-Meyer score of the upper extremity, from the acute to either of the following stages.

#### Tensor decomposition

The PARAFAC decomposition of TMS-induced oscillations in healthy adults is shown in Supplementary Fig. S1. Supplementary Table S1 shows the percentage of explained variance and the core consistency diagnosis (corcondia) obtained by decomposing the tensor with 1 to 8 components. The corcondia is a feature specifically designed for decomposition methods that indicates if the data can be fully modeled in a multilinear manner (units: %, where 100% indicates perfect multilinear data). For all the tested models, the explained variance increased in a nonlinear manner with the number of computed components, while the corcondia dropped from 100% to 0% in parallel. The final number of modes selected (3) is a good trade-off between achieving a plateau of at least 50% of explained variance, while preventing worse diagnosis regarding the proportion of variation effectively explained by a multidimensional linear phenomenon, as the corcondia value was already weak (15%) or null (Table S1). Besides, the 3 computed components were physiologically meaningful in all seven analyses, with the extra components constituting only the repetition or overlap of the three first components, as also found earlier by *(35)* (see Fig. S2). Supplementary Fig. S2 details the results of the permutation tests performed on the 4D and 5D data tensors to assess differences between stroke stages.

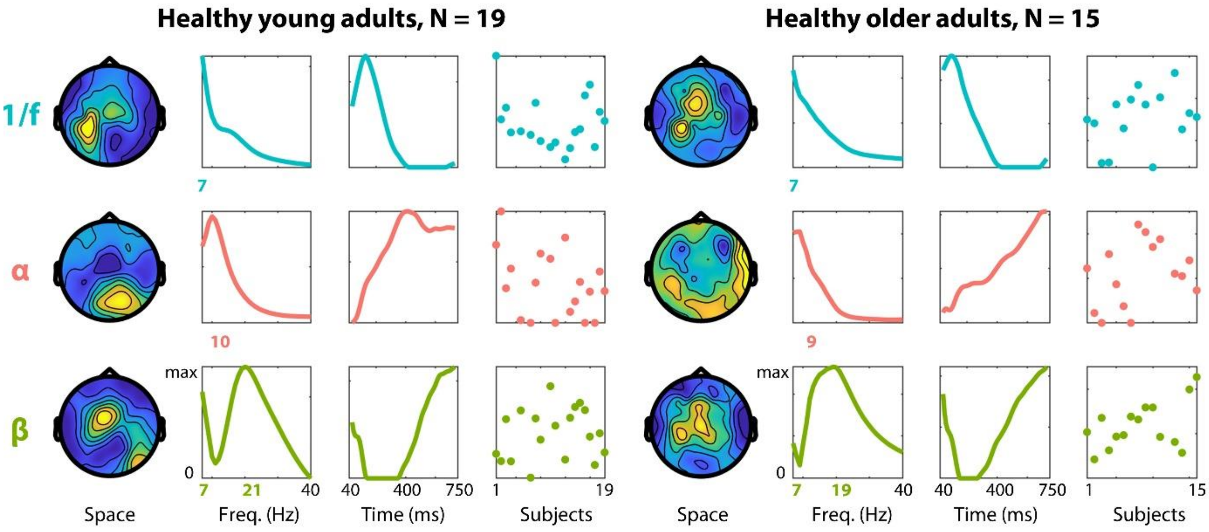


**Fig. S1. Brain oscillatory modes in healthy adults**. PARAFAC decomposition of the 4D tensor of the TMS-induced oscillations in healthy young (left) and older (right) adults. Modes are sorted by row according to their main frequency peak, from low frequencies (1/f spectral trend, blue) and α (8 Hz, red) to β (15 Hz, green) frequency bands (top to bottom). Each column depicts the relative weights (from 0 to maximum) of each mode in the space, frequency, time and patient dimensions (from left to right). The mode frequency peak is highlighted in color on the y-axis.

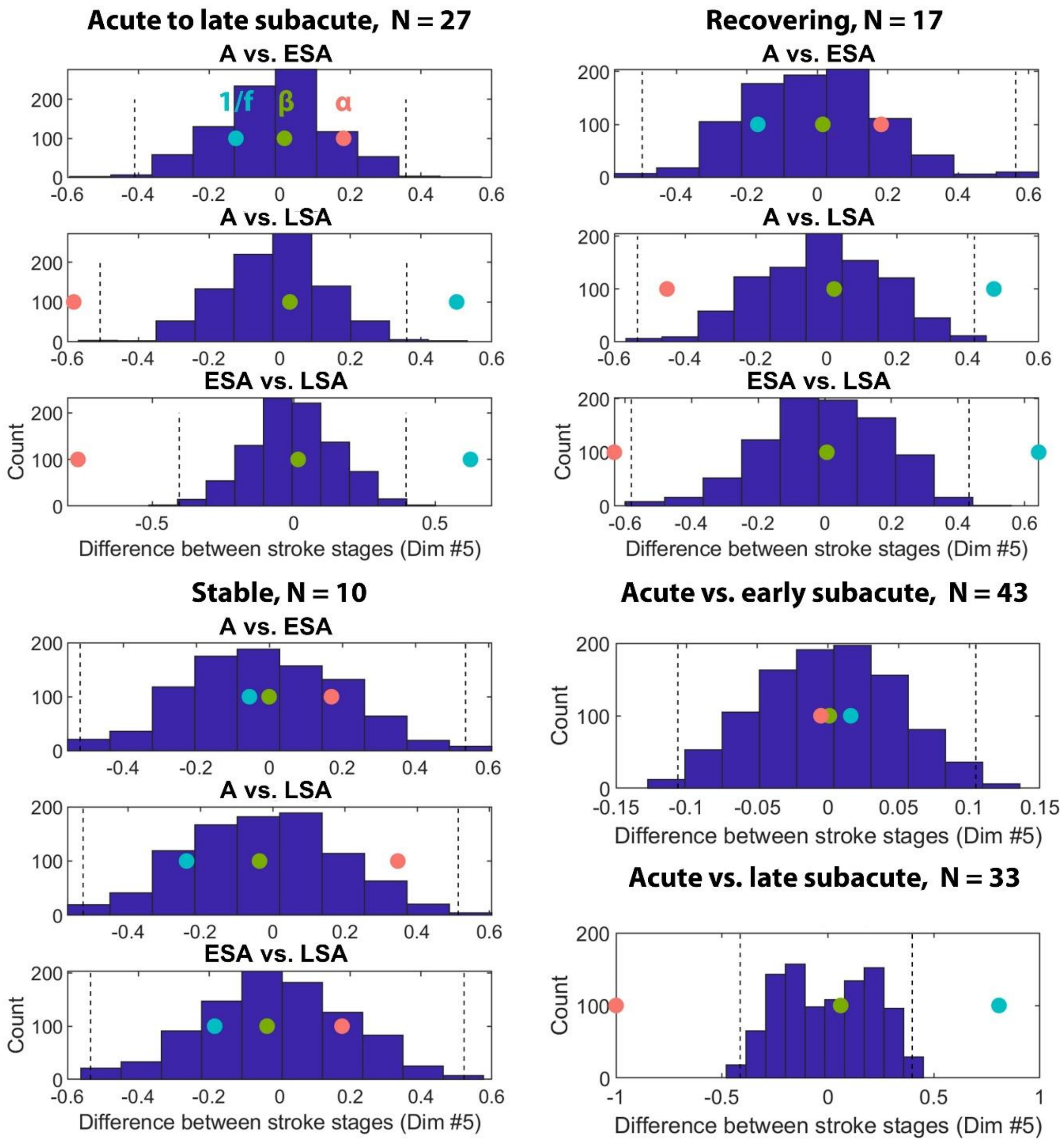


**Fig. S2. Detailed results of the permutation tests used to assess differences between stroke stages**. Each panel shows the histogram of the differences between stroke stages (over the 5$^{th}$ dimension) from 1,000 permuted surrogate tensors on the 4$^{th}$ and 5$^{th}$ dimensions (patient and stroke-stage dimensions, see *Statistical analysis*). The colored dots indicate the observed - *true* - differences obtained after the decomposition of the real data tensor, for the 1/f (blue), alpha (red) and beta (green) modes. Differences are considering significant when lower or greater than 0.83% or 0.25% of the surrogate distribution, depicted by the dashed vertical bars (Bonferroni-corrected thresholds for tensors including 2 or 3 stroke stages respectively).

| N | A stage | | Healthy young adults | | Healthy older adults | | A to LSA stages | | Recovering patients | | Stable patients | |
|---|---|---|---|---|---|---|---|---|---|---|---|---|
| | % Var | Cor. | % Var | Cor. | % Var | Cor. | % Var | Cor. | % Var | Cor. | % Var | Cor. |
| 1 | 49.9 | 100 | 45.4 | 100 | 41.1 | 100 | 47.5 | 100 | 48.1 | 100 | 47.2 | 100 |
| 2 | 53.9 | 59.9 | 53.5 | 69 | 46.7 | 32.1 | 49.6 | 1.3 | 51.8 | 37 | 50.2 | 76.5 |
| 3 | 56.8 | 15.1 | 60.3 | 15.5 | 50.4 | 4.9 | 53.5 | 0 | 55.8 | 0 | 53.7 | -1 |
| 4 | 58.7 | 0.2 | 63.7 | 0.8 | 52.5 | 0.5 | 56.2 | 0 | 58.7 | 0 | 55.0 | 0.4 |
| 5 | 60.4 | 0 | 66.4 | 0.3 | 54.2 | 0.1 | 57.7 | 0 | 60.6 | 0.1 | 58.0 | 0.1 |
| 6 | 61.7 | 0 | 69.4 | 0 | 55.7 | 0.1 | 59.2 | 0 | 62.2 | 0 | 59.4 | 0 |
| 7 | 63 | 0 | 70.8 | 0 | 57.8 | 0 | 60.6 | 0 | 63.5 | 0 | 60.7 | 0 |
| 8 | 64 | 0 | 72 | 0 | 59 | 0 | 61.4 | 0 | 64.4 | 0 | 61.7 | 0 |

**Table S1.** Explained variance (% Var) and core consistency diagnosis (corcondia – cor.) of the PARAFAC decompositions, using from 1 to 8 components (N).
